# Bromhexine Hydrochloride Prophylaxis of COVID-19 for Medical Personnel: A Randomized Open-Label Study

**DOI:** 10.1101/2021.03.03.21252855

**Authors:** Evgeny N. Mikhaylov, Tamara A. Lyubimtseva, Aleksandr D. Vakhrushev, Dmitry Stepanov, Dmitry S. Lebedev, Elena Yu. Vasilieva, Alexandra O. Konradi, Evgeny V. Shlyakhto

**Author notes:** **Address for correspondence:** Assoc. prof. Dr. Evgeny N. Mikhaylov, MD, PhD, Institute of Heart and vessels, Almazov National Medical Research Centre, 197341 Saint-Petersburg, Russian Federation. Authors ENM and TAL contributed equally to this work.

## Abstract

**Background:** Bromhexine hydrochloride has been suggested as a TMPRSS2 protease blocker that precludes the penetration of SARS-CoV-2 into cells. We aimed to assess the preventive potential of regular bromhexine hydrochloride intake for COVID-19 risk reduction in medical staff actively involved in the evaluation and treatment of patients with confirmed or suspected SARS-CoV-2 infection.

**Methods:** In a single-center randomized open-label study, medical staff managing patients with suspected and confirmed COVID-19 were enrolled and followed up for 8 weeks. The study began at the initiation of COVID-19 management in the clinic. The study was prematurely terminated after the enrollμent of 50 participants without a history of SARS-CoV-2 infection: 25 were assigned to bromhexine hydrochloride treatment (8 mg 3 times per day), and 25 were controls. The composite primary endpoint was a positive nasopharyngeal swab polymerase chain reaction (PCR) test for SARS-CoV-2 or signs of clinical infection within 28 days and at week 8. Secondary endpoints included: time from the first contact with a person with COVID-19 to the appearance of respiratory infection symptoms; the number of days before a first positive SARS-CoV-2 test; the number of asymptomatic participants with a positive nasopharyngeal swab test; the number of symptomatic COVID-19 cases; adverse events.

**Results:** The rate of the combined primary endpoint did not differ significantly between the active treatment group (2/25 [8%]) and control group (7/25 [28%]); P=0.07. A fewer number of participants developed symptomatic COVID-19 in the treatment group compared to controls (0/25 vs 5/25; P = 0.02).

**Conclusion:** Although the study was underpowered, it showed that Bromhexine hydrochloride prophylaxis was associated with a reduced rate of symptomatic COVID-19. The prophylactic treatment was not associated with a lower combined primary endpoint rate, a positive swab PCR test, or COVID-19. (ClinicalTrials.gov number, NCT04405999)

## INTRODUCTION

Severe acute respiratory syndrome coronavirus 2 (SARS-CoV-2) is the cause of respiratory disease, COVID-19 [1]. The fast SARS-CoV-2 distribution resulted in the pandemic, and the number of infected subjects keeps growing [2]. Current strategies for threat reduction include non-pharmacologic prophylactic measures, such as social distancing, masks and hand sanitizers,^2^ as well as antiviral and anti-cytokine agents [3, 4]. In recent trials, pharmacologic preventive drugs, such as anti-malaria and antibiotic medications, have failed to prevent infection, while effective prevention is urgently required [5, 6].

The SARS-CoV has been shown to utilize the endosoμal cysteine proteases cathepsin B and L (CatB/L) and the transmembrane protease serine type 2 (TMPRSS2) for binding of the S-protein [7]. It has been suggested that SARS-CoV-2 penetrates alveolar cells using the same mechanism [8]. The viral S-protein is attached to angiotensin-converting enzyme 2 (ACE2) of pneumocytes. Then it adheres to TMPRSS2 in S1 and S2-subunits [2, 7, 8, 9, 10, 11, 12]. providing the possibility for the virus to enter the cell. According to this mechanistic perspective, the protease TMPRSS2 can be targeted for preventing the penetration of SARS-CoV-2 into cells [7, 13, 14]. Two medications that have been shown to block TMPRSS2 *in vitro*, camostat mesylate and bromhexine hydrochloride; both block the ability of the protease to activate a zymogen precursor of tissue plasminogen activator. Importantly, previous studies have demonstrated that TMPRSS2 is blocked by a significantly lower concentration of bromhexine hydrochloride than required to inactivate other proteases in χell culture [8, 13].

Therefore, bromhexine hydrochloride is thought to prevent the penetration of SARS-CoV-2 into cells. If this would be confirmed in clinical settings, the drug might be used as a prophylactic medication in subjects with a high risk of infection, including medical staff. Medical staff working with COVID-19 patients are at higher risk of infection, and preventive measures, including vaccination and possible pharmacologic prevention, are of paramount importance.

We aimed to assess the preventive potential of regular bromhexine hydrochloride intake for reduction of the risk of COVID-19 in medical staff actively involved in the evaluation and treatment of patients with confirmed or suspected SARS-CoV-2 infection. The study was conducted in the period before any vaccine against COVID-19 became available.

## METHODS

### Study design

We conducted a single-centre randomized, open-label study to evaluate 8-week prophylaxis with bromhexine hydrochloride during the period of regular exposure to Covid-19 (ClinicalTrials.gov number, NCT04405999). We randomly assigned health care providers in a 1:1 ratio either to bromhexine hydrochloride or to a control group.

Study enrollment started on May 13, 2020, when the Almazov Centre began the COVID-19 treatment program. After starting the enrollment, a subsequent Federal order shortened the period of hospital repurposing, and the period for healthcare providers’ enrollment was shortened to two weeks, accordingly. Therefore, the number of enrolled subjects reached only 50 (25 subjects per group), which accounted for 42.4% of the initially estimated number. The admission of COVID-19 patients was performed between May 13, 2020 and July 25, 2020. The follow-up of the study subjects consisted was 8 weeks. The study was terminated on August 9, 2020 after reaching the end of follow-up.

The study was approved by the Almazov National Medical Research Centre ethics committee (protocol #0105-20-02C from May 12, 2020) and conducted in accordance with the Declaration of Helsinki. The study protocol is provided in the English language in **Supplements**.

### Participants

We included participants over 18 years who were employed within the emergency departments, intensive care units, and clinical departments where patients with confirmed/suspected COVID-19 were admitted. All participants were obliged to wear personal protective equipment (PPE) as prescribed by WHO recommendations and local instructions. The PPE included respirators class FFP2 or FFP3, full skin covering, and protective eyeglasses. Blood tests for SARS-CoV-2 antibodies were taken at baseline before randomization (serologic qualitative assessment of IgM and IgG, ELISA-BEST, Vektor-Best, Novosibirsk, Russia).

Participants were excluded if they had symptoms of respiratory infection within the last 2 months or a history of COVID-19, or a positive nasopharyngeal swab polymerase chain reaction (PCR) test to SARS-CoV-2 before the day of randomization, χonfirμed direct contact to a subject positive for SARS-CoV-2 within the last 14 days or had a positive serologic test (either IgM or IgG). The additional exclusion criteria were a history of gastric ulcer or other contraindications to bromhexine hydrochloride, pregnancy, and any severe chronic disease.

### Setting

Recruitment was performed via the institution’s electronic communication systeμ and via personal contacts with healthcare providers. The participants provided a scanned copy of the signed consent. We performed follow-up phone calls and sent e-mail surveys on days 7, 14, 21, 28, and at week 8. The survey asked about any follow-up testing, illness, or hospitalizations. Participants who did not respond to follow-up surveys were actively contacted by text messages and telephone calls.

### Interventions

The study statisticians adjusted a software for randomization using the minimization method weighing the following factors: age (categories: 18-45, 45-64, 65-74, 75-79 years), the type of anticipated contacts with SARS-CoV-2 (confirmed SARS-CoV-2 infection cases within the “red” zone, working closely with colleagues who had proven contacts with SARS-CoV-2 patients). A study coordinator sequentially assigned participants. The assignments were open to investigators and participants.

Bromhexine hydrochloride was dispensed and shipped to participants by the study coordinator. The dosing regimen for oral bromhexine hydrochloride was 8 mg 3 times per day starting from the day before the first contact with COVID-19 (first day of work in a repurposed department). The dosing regimen was based on previous reports [8, 15].

The control group was not prescribed any additional drug or placebo.

### Outcomes

The composite primary endpoint was prespecified as a positive nasopharyngeal swab SARS-CoV-2 PCR test or the presence of clinical symptoms of infection within 8 weeks after starting contacts with COVID-19 subjects. COVID-19–related symptoms were assessed based on the U.S. Council for State and Territorial Epidemiologists Criteria for confirmed cases (positive nasopharyngeal swab PCR test), probable cases (presenχe of cough, shortness of breath, or difficult breathing, or the presence of two or more symptoms of fever, chills, rigors, myalgia, headache, sore throat, and new olfactory and taste disorders), and possible cases (the presence of one or more compatible symptoms, which could include diarrhea) [16].

Secondary endpoints included: time from the first contact with a person with suspected/confirmed COVID-19 to the appearance of respiratory infection symptoms; the number of days before the first positive SARS-CoV-2 test; the number of asymptomatic participants with a positive nasopharyngeal swab test; the number of mild, moderate and severe COVID-19 cases; adverse events possibly related to bromhexine hydrochloride.

According to the study protocol, nasopharyngeal swab SARS-CoV-2 PCR tests were performed every 7 days, and additional tests were performed in case of infection. The PCR test was performed by qualitative analysis (DNA-technology, Moscow, Russia).

Outcomes were measured at 7, 14, and 28 days after study enrollment, and then at 8 weeks. Outcome data including PCR, COVID-19–related symptoms, adherence to the study drug, and side effects, as well as not taking the study drug by the control group subjects were collected through participants’ reports.

### Sample size

Initially, we estimated that 59 subjects would need to be enrolled in each group (140 subjects in total, with an assumed 15% drop-out rate), when calculated using an alpha of 0.05 and 80% power, assuming a primary endpoint achievement in 10% of treatment group subjects, and 30% in the control arm. These numbers were derived from previous reports on the incidence of COVID-19 positivity in first-line health care workers, and reports demonstrating a higher proportion of COVID-19 positive Russian physicians [17-19]. However, as described in the Methods section, the enrollment was ceased when 50 subjects (25 per group), were included. The number of included subjects was amended by the study committee and updated on the clinicaltrials.gov website after study completion on September 3, 2020.

### Statistical analysis

The primary and secondary endpoints were assessed with the Mann-Whitney U test. Statistical analysis was performed using STATISTICA software 13.0 (StatSoft, USA), according to the intention-to-treat principle, with a P-value suggesting a statistically significant difference when <0.05.

## RESULTS

### Participants

Following a general notification via an institutional electronic system, one hundred and fifty healthcare providers were contacted personally, and 62 positive responses were obtained. Among 62 subjects who agreed to participate, 50 persons fulfilled the inclusion criteria (80.6%) (**Figure 1**).

**Figure 1.**
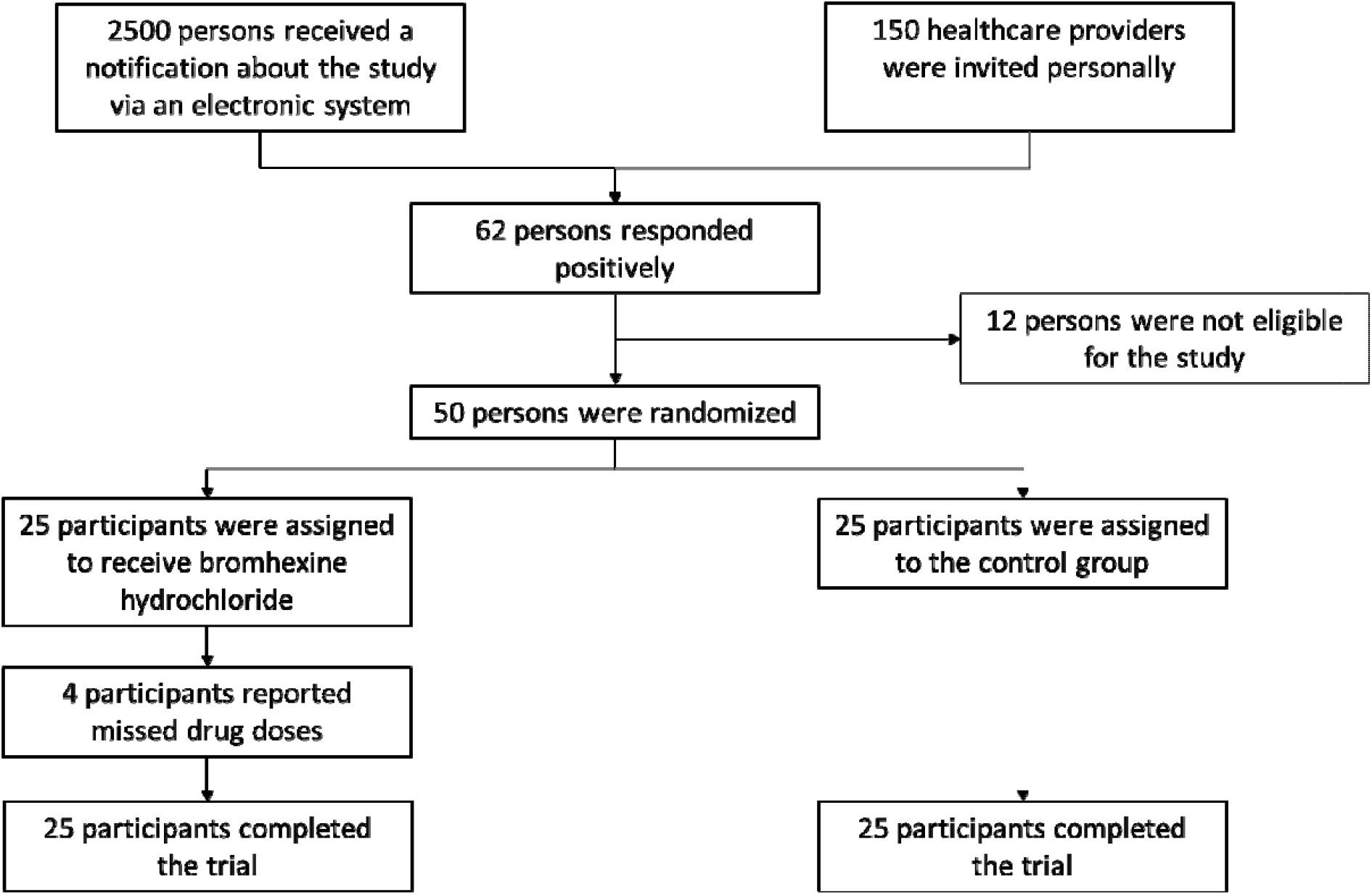
Study flowchart.

The demographic and clinical characteristics of the participants are presented in **Table 1**. Three (6%) of the participants were hypertensive, 2 participants had hypercholesterolemia. Physicians comprised 88% of the population, and nurses 12%. The median exposure time to patients with confirmed/suspected COVID-19 was 7.5 hours per week.

**Table 1.**
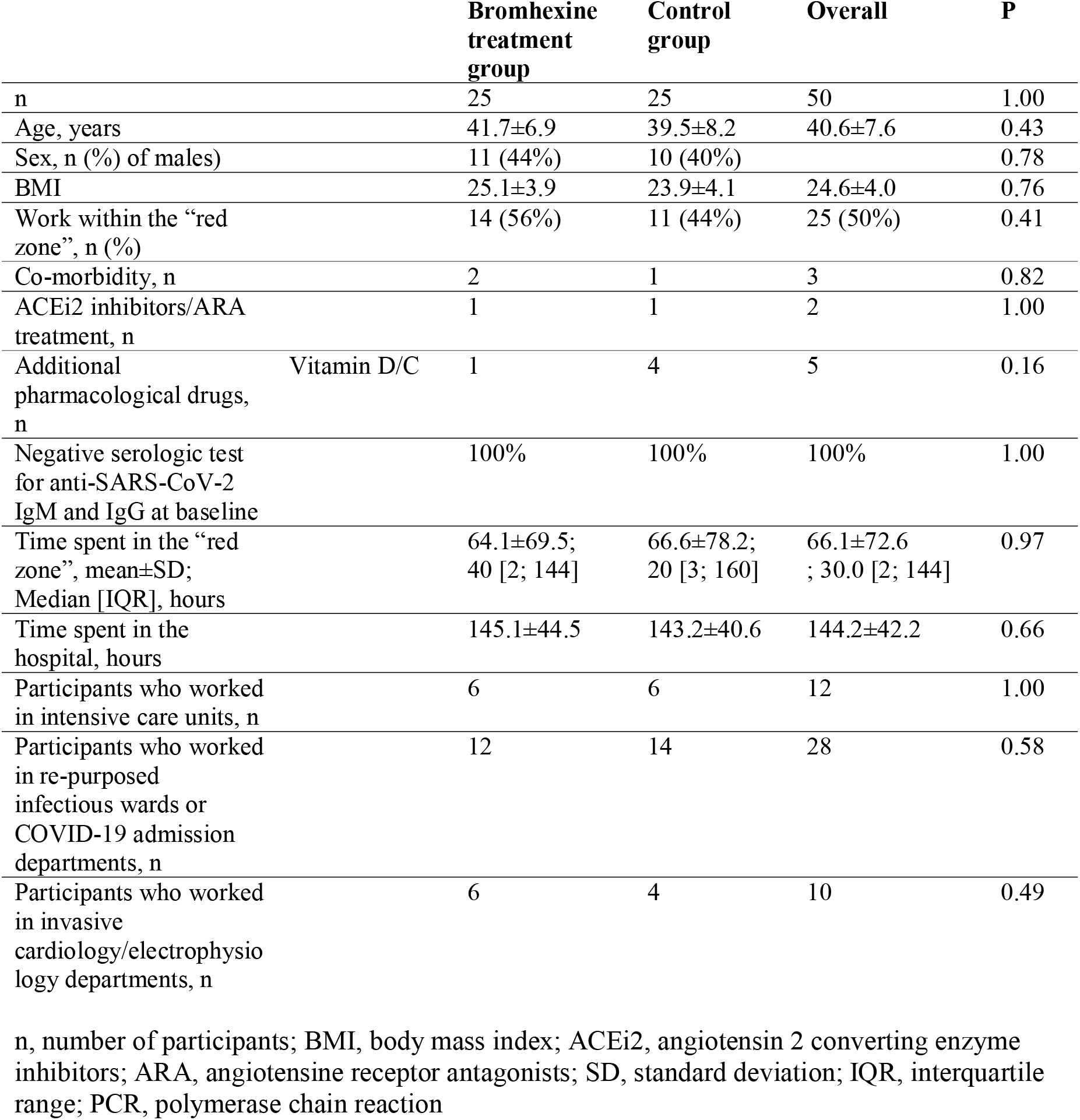
Demographic, clinical and work characteristics of study participants.

Overall, 80% (40 out of 50) participants had a very high-risk exposure (contacting with aerosols from intubated COVID-19 patients) and 20% (10 out of 50) - a relatively lower risk exposure (contacting with suspected/confirmed COVID-19 patients). All the participants had 6-hour duration shifts working with COVID-19 patients, used FFE respirators, eyeglasses, and skin coverings, and had direct contact with staff outside the “red zone” without PPE.

Seven (14%) participants used vitamins D, C as an intention to prevent infeχtion: 3 participants in the bromhexine group (3/25, 12%) and 4 participants in the control group (4/25, 16%; P>0.05). None of the participants reported any unprotected contact with COVID-19 patients outside the hospital.

There were no subject losses nor dropouts after randomization.

### Primary endpoint

All participants completed all scheduled surveys. The primary endpoint, a positive swab test and/or infection symptoms, was documented in 9/50 (18%) participants during the follow-up period: in 8 - by day 28, and in 1 - by week 8 (**Tables 1 and 2**). The primary outcome rate did not differ significantly between the treatment and control groups (2/25 (8%) vs 7/25 (28%), respectively) (P=0.07). Two hospitalizations for COVID-19 pneumonia were reported in the control group (2/25, 8%), and none – in the bromhexine group (0/25, P=0.16). No severe cases with intensive care unit adμission or death occurred.

**Table 2.** Study outcomes: positive PCR tests and COVID-19.

|  |  | **Bromhexine**<br>**treatment**<br>**group** | **Control**<br>**group** | **Overall** | **P** |
| --- | --- | --- | --- | --- | --- |
| n |  | 25 | 25 | 50 | 1.00 |
| Primary outcome<br>(Positive PCR test or<br>COVID-19) |  | 2 | 7 | 9 | 0.07 |
| Positive SARS-CoV-2<br>PCR test, n |  | 2 | 7 | 9 | 0.07 |
|  | Asymptomatic | 2 | 2 | 4 | 1.00 |
|  | Symptomatic | 0 | 5 | 5 | 0.02 |
| COVID-19 moderate, n |  | 0 | 3 | 3 | 0.08 |
| COVID-19 severe with<br>hospitalization, n |  | 0 | 2 | 2 | 0.16 |
| Drug non-adherence |  |  |  |  |  |
|  | Missed doses<br>of<br>bromhexine, n | 4 | - | - | - |
|  | Number of<br>missed<br>bromhexine<br>doses, Median<br>[IQR] | 6 [5.25; 7.5] | - | - | - |
| Adverse events, n |  | 2 | 0 | 2 | 0.16 |

n, number of participants; IQR, interquartile range; PCR, polymerase chain reaction

### Secondary endpoints

There was no difference between the groups in the time between the first contact with a suspected/confirmed COVID-19 person and infection symptoms, or an asymptomatic positive PCR test. The first positive SARS-CoV-2 PCR tests in the treatment group appeared at 2 and 3 weeks, and in the control group the positive tests were obtained at 2, 3, 4 weeks, and during weeks 5-8 (**Table 3**). Both SARS-CoV-2 PCR positive participants in the bromhexine treatment group continued the intake of the drug until two consecutive negative tests.

**Table 3.** Timeline of positive SARS-CoV-2 tests in both groups.

|  |  | **Days 1-7** | **Days 8-14** | **Days 15-21** | **Days 22-28** | **Weeks 5-8** | **Total** |
| --- | --- | --- | --- | --- | --- | --- | --- |
| Number of participants with a positive PCR test | Treatment group | 0/25 | 1/25 | 1/24 | 0/23 | 0/23 | 2/25 |
|  | Control group | 0/25 | 2/25 | 3/23 | 1/20 | 1/19 | 7/25 |
| Number of participants with clinical symptoms of respiratory infection | Treatment group | 0/25 | 0/25 | 0/25 | 0/25 | 0/25 | 0/25 |
|  | Control group | 0/25 | 0/25 | 2/25 | 3/23 | 0/20 | 5/25 |

Numbers of subjects with an outcome are divided by the number of subjects at risk. Three patients developed clinical symptoms after positive PCR test in the previous period (days 8-14 and days 15-21). $P > 0.05$ for each period of follow-up.

Among 9 persons with a positive PCR test, 4 were asymptomatic (2/25 – in the treatment group and 2/25 – in the control group; P>0.05). Five participants from the control group had infection symptoms: only respiratory virus infection symptoms - in 3 persons; pneumonia – in 2. All participants with respiratory symptoms were positive for SARC-Cov-2 and were compatible with confirmed COVID-19 per the U.S. case definition.

Taking together, symptomatic COVID-19 was diagnosed in the control group only: no cases in the treatment group (0/25) vs 5/25 cases in the control group, P=0.02.

### Adherence and safety

Missed days of bromhexine intake were reported by 4 participants, the duration of skipped treatment was 1-4 days. One participant who missed two days of drug intake had a positive SARS-CoV-2 PCR test (asymptomatic case). It was impossible to assess whether missed drug intake and infection occurred on the same day.

Two participants reported adverse events possibly related to bromhexine hydrochloride treatment: a short period of hot flashes at treatμent initiation and transient cough. No cases of treatment termination or interruption due to adverse events were reported. There were no adverse events in the control group. No statistically significant difference was found in the rates of adverse events between the groups.

No subjects in the control group reported the intake of the study medication.

## DISCUSSION

Although generally underpowered, the present study has several important findings. The primary combined endpoint, the rate of positive nasopharyngeal swab PCR tests for SARS-CoV-2 or symptomatic COVID-19, was similar in both groups. However, there was a trend towards a lower rate of the positive swab PCR test in the bromhexine hydrochloride treatment group. Importantly, the rate of clinically significant SARS-CoV-2 infection was statistically lower in the treatment group (0/25 participants) compared with the control group (5/25 participants).

Even though the enrollment was terminated prematurely, it should be acknowledged that the rates of the primary outcome in the active treatment and control groups were equal to the estimated initially (8% vs 10%, and 28% vs 30%, respectively). This suggests that the primary hypothesis of a 20% reduction in the outcome might be possible with the prevention treatment. Indeed, the main outcome presents an important numerical difference but without reaching statistical significance, and should be interpreted with consideration of the limited sample size.

The risk of symptomatic and severe COVID-19 depends on age and comorbidity [20]. Our study population was generally younger and healthier than in the majority of COVID-19 studies. However, in a study of post-exposure COVID-19 prophylaxis by hydroxychloroquine, the mean age and average health status of the participants were comparable [5]. Thus, for prevention efficacy assessment, our group can be informative. Moreover, the fact that the safe and cheap drug showed the risk reduction of symptomatic infection can be important for future larger studies and practical use.

We used a thorough approach for detecting asymptomatic cases performing nasopharyngeal swab PCR tests every week of follow-up using the approved methodology [21].

Two factors increase the value of our study: good adherence, which is significantly higher, compared to generally published prevention studies, and the high survey response rate. This can be explained by the high motivation and high awareness of the medical staff.

It should be mentioned that a small number of participants used additional medications, including ACE2 inhibitors/ARA, or vitamin C, D. No statistical difference in the use of additional pharmacological treatment was documented between the groups, and the number of subjects with additional medications was very small, thus we can conclude that this aspect had no impact on the study results. It is plausible that the results may be more robust with a placebo-controlled group. Due to the organizational difficulties during a limited period of study preparation, the provision of placebo was impossible.

Although all healthcare workers were instructed to wear protective masks and medical gloves outside the hospital, we cannot exclude that some of the participants might have been infected outside the hospital or during direct communication within the community during rest and meals. However, if bromhexine hydrochloride prevention is effective, it should be effectiveanywhere.

It is generally believed that the risk of infection is lower when the staff uses PPE as appropriate. Thus, further reduction of clinically significant cases can be an argument for the implementation of this approach into clinical practice, even before obtaining better evidence.

In a recently published randomized study of bromhexine hydrochloride treatment in patients hospitalized with COVID-19, the authors have shown that the early administration of oral bromhexine reduces the intensive care unit transfer, intubation, and mortality rate [15]. In another small open-label randomized study, where 18 patients with COVID-19 were included, bromhexine hydrochloride treatment has been shown associated with a trend towards the improvement in chest computed tomography, the need for oxygen therapy, and discharge rate within 20 days [22]. In this study, a much higher drug dosage was utilized (32 mg t.i.d.). Of note, subjects were patients with confirmed mild or moderate COVID-19 disease, while in our study healthy subjects were given the medication for prophylactic purposes. We have chosen a lower dose of the drug (8 mg t.i.d.) according to previous research demonstrating its efficacy [8, 15]. A higher Bromhexine hydrochloride dose might be associated with a higher rate of adverse events, but this assumption is speculative. Although hypothesis regarding the positive effects of bromhexine on viral penetration into cells in noninfected subjects differs from the background presented in the two latter studies, these investigations support the assumption of positive effects of the drug against SARS-CoV-2.

Whether the prophylactic use of bromhexine hydrochloride against SARS-CoV-2 infection in the general population is effective is a separate question that should be answered in larger-scale randomized trials.

### Study limitations

The major limitation of the present study is the small number of participants. The initial estimation of the sample size was higher, but the enrollment was ceased earlier due to the reasons declared in the Methods section. On the other hand, the sample size was sufficient to demonstrate the significant difference in secondary outcomes between the groups.

Another limitation is the lack of sequential serologic testing, which we considered less informative due to a very short study period.

It should be mentioned that safety and side effects data from self-reports is probably limited. Theoretically, there was a risk of crossover between the groups; however, no evidence of at least one crossover was found.

Additionally, there was no objective supervision of how and where the drugs were taken, only weekly surveys were distributed and collected from participants via electronic facilities; therefore, we were not able to provide any direct observation of the therapy, and the validity of the intervention could not be completely verifiable.

In contrast to the previous study by Li T et al [22], we did not assess potential effects of Bromhexine hydrochloride on COVID-19-related lung or hepatic injury, since the percentage of patients with overt disease was small.

## CONCLUSIONS

In this randomized single-centre open-label study, bromhexine hydrochloride prophylactic treatment was associated with a reduction in the rate of symptomatic COVID-19 among medical personnel. However, the primary combined outcome, the rate of a positive nasopharyngeal swab PCR test and/or COVID-19, did not reach a statically significant difference.

Although generally underpowered, this study suggests that bromhexine hydrochloride may offer clinical value when taken as a prophylactic treatment.

## Data Availability

All date are available upon request to the authors

## ACKNOWLEDGEMENTS

The authors thank all study participants for their dedication to participate and adherence to the study protocol. We thank Dr. Olesya V. Melnik, MD, PhD, for her invaluable help in serologic data management. The research was performed using the core facilities of Biobank of the Almazov National Medical Centre.

A preprint of this manuscript has previously been published [23].

## AUTHOR CONTRIBUTIONS

Research idea: ENM, TAL, DS. Study design: ENM, TAL, AOK, EVSh. Data collection and management: ENM, TAL, ADV, DSL, EYuV. Data interpretation and analysis: ENM, TAL, ADV, DS, DSL, AOK, EVSh. Writing the artiχle draft: ENM, TAL, AOK. Critical review and approval of the article final version: all authors.

## STUDY HIGHLIGHTS

- Bromhexine hydrochloride is a TMPRSS2 protease blocker that is thought to preclude the penetration of SARS-CoV-2 into cells;
- Prophylactic treatment with bromhexine hydrochloride reduces the rate of clinical infection among medical workers who manage patients with COVID-19

**SUPPLEMENT FILE**. This file contains study protocol translated to English.

